# Fast clinical trial identification using fuzzy-search elastic searches: retrospective validation with high-quality Cochrane benchmark

**DOI:** 10.1101/2023.09.06.23295135

**Authors:** Willem M Otte, David G P van IJzendoorn, Philippe C Habets, Christiaan H Vinkers

## Abstract

The synthesis of treatment effects relies on systematic reviews of intervention trials. This process is often laborious due to the need for precise search queries and manual study identification. Recent advancements in database architecture and natural language processing (NLP) offer a potential solution by enabling faster, more flexible searches and automated extraction of information from unstructured texts.

Our study assesses the effectiveness of NLP-based literature searches within a novel database structure in comparison to the Cochrane Database of Systematic Reviews. We created a user-friendly elastic search database containing 36 million PubMed-indexed entries. We developed reliable filters for identifying randomized clinical trials and clinical intervention studies, as well as extracting relevant subtext related to population and intervention.

Our results indicate a high precision of 0.74, recall of 0.81, and F1-score of 0.77 for population subtext, and a precision of 0.70, recall of 0.71, and an F1-score of 0.70 for intervention subtext. Our approach efficiently identified included studies in 90% of systematic reviews, missing no more than two trials compared to Cochrane. Furthermore, it produced fewer total hits than a comparable PubMed keyword search, demonstrating the potential of the new database structure to enhance the efficiency and effectiveness of aggregating clinical evidence.

## 1. Introduction

Evidence-based medicine requires synthesizing the latest and most valid estimates of clinical intervention effects. As a result, systematic reviews with meta-analyses of randomized controlled trials and other clinical intervention designs are regularly published in journals, including the Cochrane Database of Systematic Reviews (CDSR) journal.^1^

Conducting a systematic review using medical databases requires careful design of sensitive search queries, which is currently the modus operandi to identify all relevant studies and simultaneously exclude irrelevant publications. The estimated labor effort to produce a systematic review ranges from six months with a single person working on the review for ten to twenty hours a week (Michelson and Reuter, 2019) to an average of sixteen months involving multiple persons (Borah et al., 2017). The time-consuming searches add to the research waste associated with systematic reviews (Roberts and Ker, 2015). A Cochrane systematic review typically requires one to two years to complete. This time and effort limits the speed and flexibility that can advance evidence-based medicine, resulting in many meta-analyses and systematic reviews on the same topic that are sometimes redundant (Chapelle et al., 2021; Puljak et al., 2023).

The reason for the duration and efforts to arrive at systematic reviews and meta-analyses is that these searches are complex and require extensive manual work that spans several databases. First, reliable and standardized meta-information and the unstructured information on the patient population and type of intervention ‘hidden’ within the larger summary text is lacking, making manual identification of database entries the cornerstone (Ossom Williamson and Minter, 2019). Moreover, databases such as MEDLINE and its interface PubMed require carefully composed keyword sets and Boolean operators. Additionally, queries often do not allow approximate or fuzzy formulation, which requires the user to spell all medical terms correctly and explicitly deal with US and UK spelling alternatives (e.g., ‘randomised’ vs. ‘randomized’, ‘anaemia’ vs. ‘anemia’, ‘tumour’ vs. ‘tumor’, ‘fibre’ vs. ‘fiber’, and ‘leucocyte’ vs. ‘leukocyte’).

Recently, new database architectures have emerged, allowing fast and fuzzy searches using non-Boolean queries (Cohen et al., 2020; Manhaeve et al., 2021). In addition, natural language processing (NLP) allows meta-information generation required to bring evidence-based medicine towards a more automated information processing. These developments enable the computerized transformation of unstructured texts into structured textual features and facilitate tasks such as extracting phrases related to clinically relevant information domains, including Population (i.e., which disease or patient’s condition is central) and Intervention (i.e., which drug or intervention is used). Combining a new database architecture with NLP meta-information generation may be a promising avenue to keep track of the latest developments in clinical literature and mitigate the need for manual labor-intensive updating. However, the quality of this approach is important. Fuzzy searching and automatic generation of meta-information can be helpful, but only if they identify the relevant clinical entries with sufficient sensitivity.

This study, therefore, aimed to compare NLP-based literature searches within a new database structure to the yield of CDSR systematic reviews that are currently the gold standard. To this end, we focused on the MEDLINE database, one of the major medical databases, containing 36 million entries from 5,200 worldwide journals covering biomedical research publications from 1966 onwards.^2^ To this end, we *i*) built a stand-alone, freely available, fuzzy-enabled elastic search database containing all 36 million PubMed-indexed entries, *ii*) constructed and validated reliable filters to tag randomized clinical trials and other clinical intervention studies, *iii*) developed and validated the extraction of Population and Intervention-relevant subtext from abstracts, and *iv*) investigated to what extent short, user-friendly and approximate queries identify the list of included studies within a random set of CDSR systematic intervention reviews.

## 2. Methods

Figure 1 provides a schematic overview of the proposed strategy used in this paper. Rather than searching PubMed directly, a new database and interface bridges the clinical studies and the user. The construction and validation are described below.

**Figure 1.**
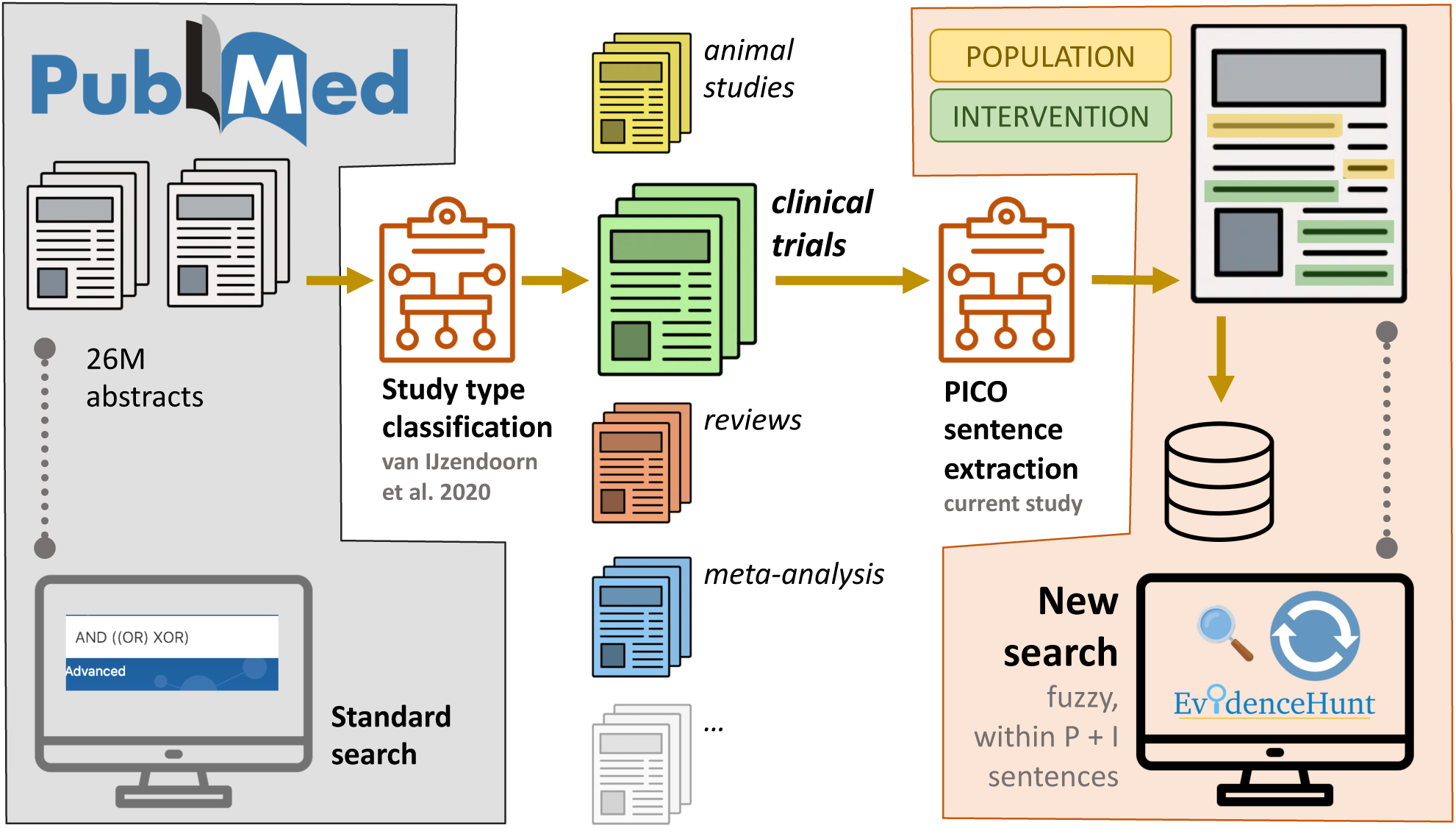
Infographics of implemented and validated study identification pipeline. Standard PubMed searches (left) require dedicated query compilation with correct spelling and are applied to 26 million abstracts. The new search (right) allows for spelling errors or term variations and is applied to pre-selected study types and extracted sentences with Population and Intervention information. The interface is available at: https://evidencehunt.com/browse/.

### 2.1 Medline Database

Once a year, a complete baseline snapshot of PubMed data is published in XML format for download.^3^ Additional updates are posted every day, including new, revised, and deleted entries.^4^ We downloaded, parsed, and incorporated this data in a custom elastic search database, continuously updated the data daily, and provided access through an open web interface.^5^ At the time of writing (Sep 2023), 35.9 million PubMed entries are indexed.^6^ Next, we built an Elasticsearch database based on all PubMed entries available. Elasticsearch provides a near real-time scalable search, indexing numbers, dates, abbreviations, diverse sets of coordinates, and almost any datatype while supporting multiple languages.^7^

### 2.2. Study type identification

We developed and validated the database entries according to the following study types: ‘randomized trial’, ‘clinical intervention’, ‘meta-analysis’, ‘systematic review’, ‘protocol’, ‘rodent study’, or ‘other’ (van IJzendoorn et al., 2022). In short, we used an active learning strategy and multiple raters to create a large training set of 50,000 PubMed abstracts. The filter performance on unseen external data was characterized by excellent sensitivities for the identification of ‘randomized trial’ studies (0.80, 95% confidence interval (CI): 0.67–0.92) and ‘clinical intervention’ studies (0.94, CI: 0.86–1.0) and specificities: 0.995 (CI: 0.99–1.0) for ‘randomized trial’ and 0.82, (CI: 0.80–0.85) for ‘clinical intervention’. The model achieved an average sensitivity and specificity of 0.94 and 0.96, respectively, on an external dataset of 5,000 abstracts. In contrast, PubMed’s internal filters had a much lower sensitivity for both systematic reviews with meta-analysis (i.e., 0.18) and randomized controlled trials (i.e., 0.26).

### 2.3 Patient and Intervention Subtext; Online interface

After identifying interventional database entries, we may focus on the text itself. Unfortunately, PubMed abstracts summarizing trials do not follow a standardized format. Therefore, a text model identifies the required patient population and intervention information.

Population and Intervention are widely used text fields within the Patient-Intervention-Control-Outcome (PICO) framework used in evidence-based medicine formulations and standardization protocols (Richardson et al., 1995). We developed a text classification model to identify the words and sentences associated with Population and Intervention in every PubMed abstract. An independent physician identified all words related to the patient Population and Intervention in 1,360 random sampled ‘randomized trial’ and ‘clinical intervention’ entry abstracts within the open-source text annotation tool Doccano.^8^ The authors checked the labels.

The annotated abstract text was converted into an Inside-Outside-Beginning (IOB) tagging scheme and randomly divided into a train (sample size: 1020), validation (170), and test (170) dataset. Next, we finetuned a PubMedBERT Named-Entity Recognition word classification model on the training set and searched for the optimal hyperparameters using the validation set. We finetuned the model for seven epochs with a learning rate of 0.0001, 400 warm-up steps, and a batch size of sixteen with the pre-trained weights of the PubMedBERT ‘base’ (abstract-only) network architecture as a starting point.^9^ The independent test set characterized the performance in terms of precision, recall, and F1-Score evaluation metrics. Precision measures how many annotated words are correctly identified (true positives). Recall estimates how many annotated words are correctly identified relative to the total number of annotated words in the abstract (sensitivity). F1-Score averages precision and recall to provide a balanced estimate if annotated word prevalence is low.

We combined the study identification, Population, and Intervention subtext extraction with an online interface, allowing flexible text input. The interface is available at: https://evidencehunt.com/browse/, and searching within ‘randomized trial’ and ‘clinical intervention’ entries is facilitated in the interface’s search fields by distinguishing three types of query building blocks suitable for different combinations of words: ‘all the words’, ‘at least one of the words’, and ‘without the words’. The building blocks are available for subtext identified as belonging to descriptive information of clinical ‘Population’ (P) or ‘Intervention’ (I).

### 2.4 Validation: Cochrane review selection

To validate the usefulness of our approach, which integrates a newly structured NLP-based database with validated study types and approximate search of Population and Intervention into an online platform, we sampled reviews from the systematic reviews in the CDSR published in their first 2022 Issue (April 4). For that, we used the NCBI’s EUtils API with the following query: “Cochrane Database Syst Rev”[journal] AND (“2022/01/01”[PDAT]: “2022/12/31”[PDAT]). Studies were ordered on publication date, and the fifty most recent reviews were selected.

We downloaded the full text and determined which reviews were dedicated to answering an interventional research question. Diagnostic, prognostic, scoping, and mixed-methods CDSR reviews were excluded. We manually identified all studies included in the systematic review analysis, also published in PubMed. We could not extract the number of PubMed studies identified within the Cochrane systematic review’s inclusion process, as these inclusions combined multiple database searches with manual reference screening. As a comparison between the number of identified studies in our system and conventional queries, we obtained the number of hits if the P and I keywords were searched in PubMed, combined with Cochrane’s latest Search Strategy recommendation.^10^ Keywords were included using their MeSH equivalent and appearance in the title or abstract.

Based on the title and abstract of systematic reviews, we constructed short and user-friendly queries compatible with the new interface layout. For example, the query for the systematic review ‘Interleukin-6 blocking agents for treating COVID-19: a living systematic review’ only included ‘COVID-19’ and ‘interleukin blocking’ as query texts. **Table 1** provides the systematic reviews and design queries. We identified all ‘randomized trial’ and ‘clinical intervention’ entries with these short queries and determined the overlap with the actually included studies in the reviews.

**Table 1.**
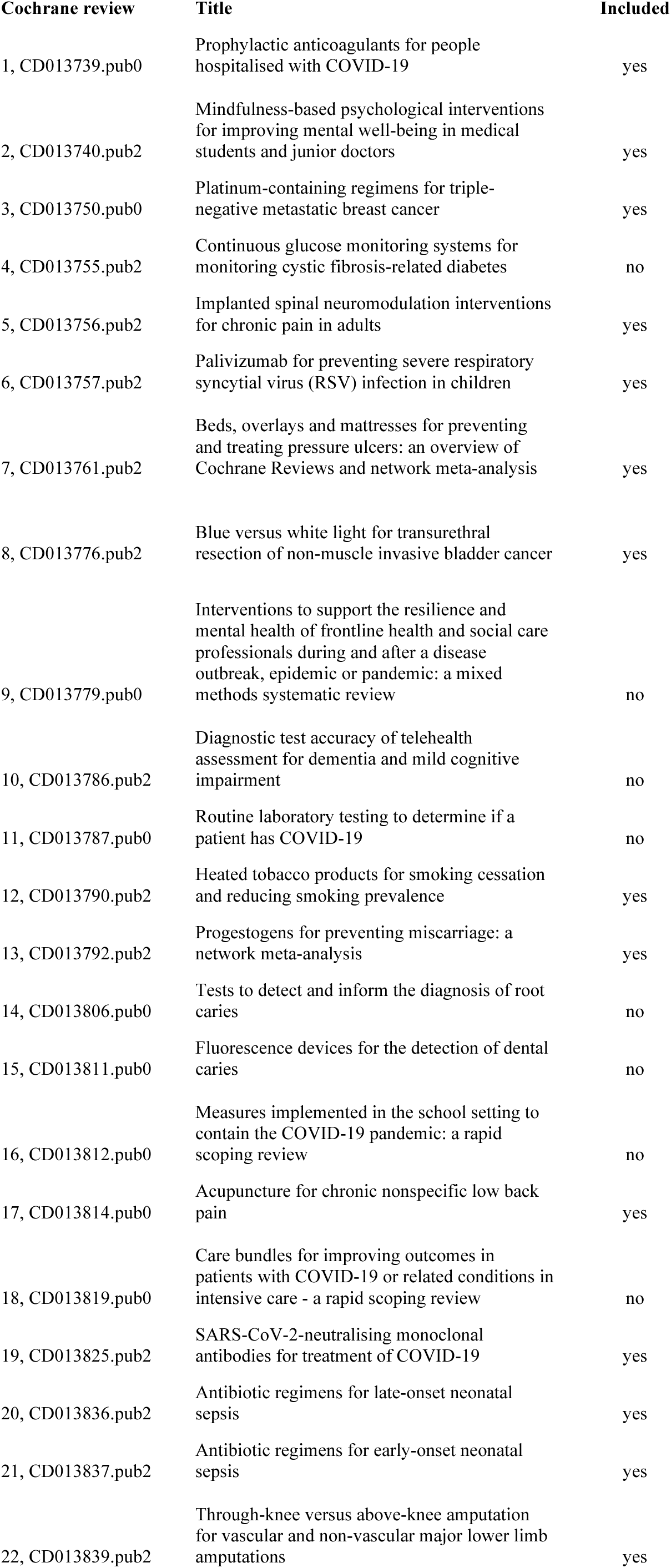
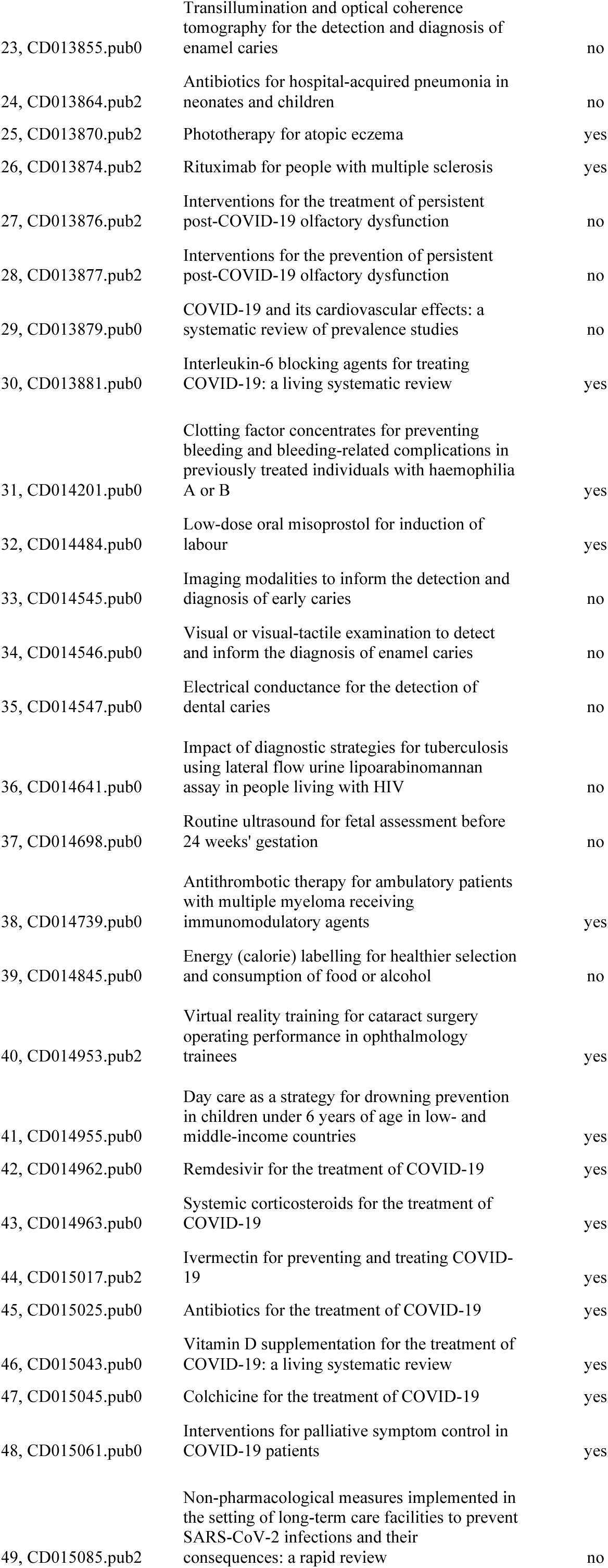
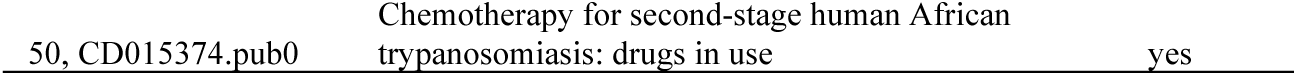
Overview of the fifty sampled systematic Cochrane reviews.

## 3. Results

In the text classification model, words associated with Population were identified with a precision of 0.74, recall of 0.81, and F1-score of 0.77. Words related to Intervention with a precision of 0.70, recall of 0.71, and an F1-score of 0.70. See Figure 2 for a representative example of classified texts on an unseen ‘randomized trial’ database entry.

**Figure 2.**
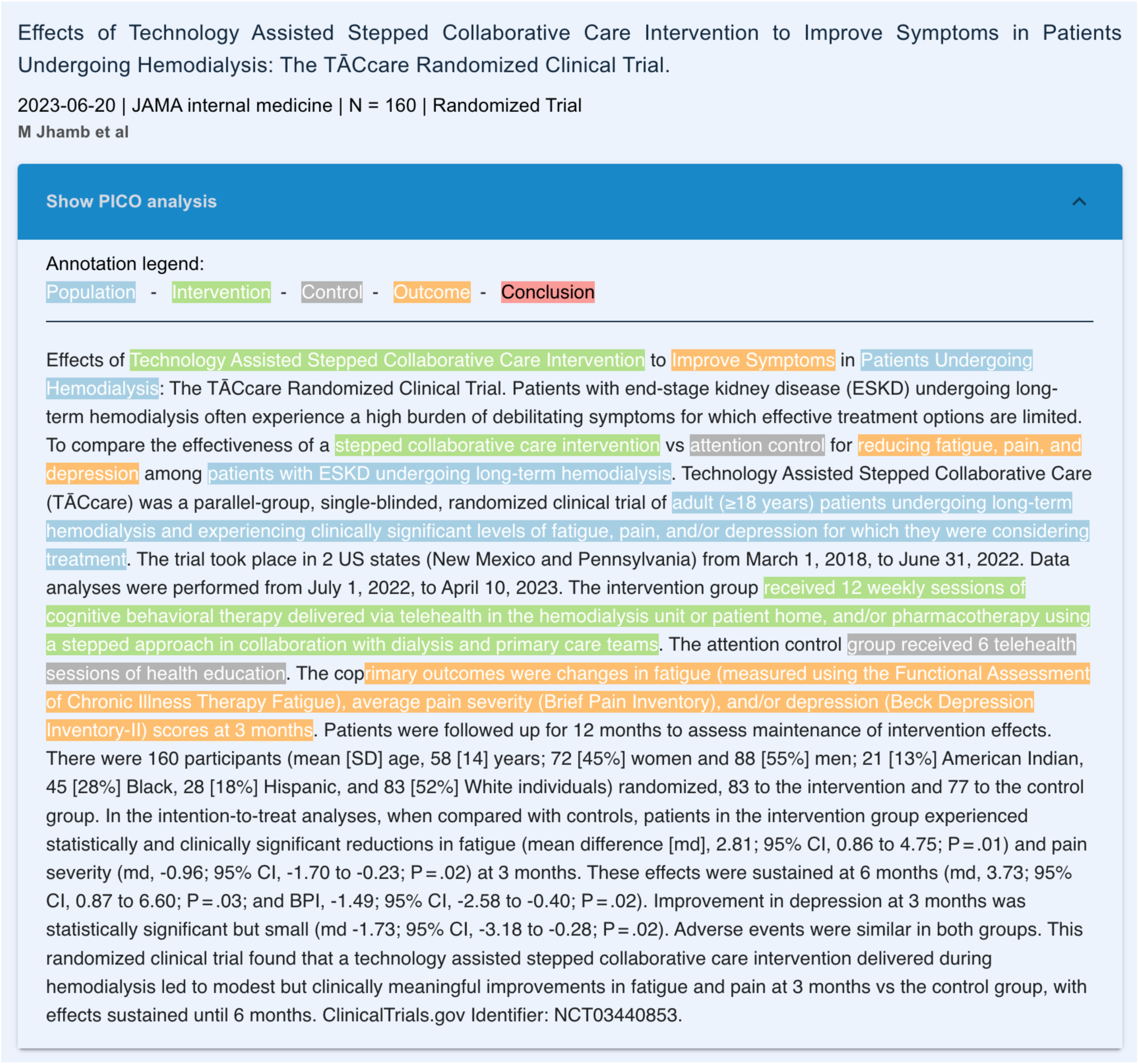
Representative example of a recent randomized clinical trial published on PubMed and imported in our elastic search database. Subtexts are automatically classified into different domains, including Population and Intervention. Queries are restricted to the Population and Intervention subtexts, ignoring the abstracts remaining.

Thirty of the fifty selected Cochrane reviews were included for validation analysis (**Table 2**). Queries were short, with a maximum length of 21 words (median: four). In eleven (37%) systematic reviews, all of the included trials were also detected with the new search strategy, and in twenty-five systematic reviews (83%), more than seventy-five percent of trials were found. In a small subset, no trials were identified. In 90% of systematic reviews (27/30), the new search strategy missed no more than two of all included trials by Cochrane. Reasons for missing trials were: study type classification ‘other’ (rather than ‘clinical trial’ or ‘human intervention’), incorrect Population or Intervention sentence extraction, or a lack of information in the abstract (e.g., empty abstract).

**Table 2.** Identified trials for the intervention systematic review, based on the Population and Intervention query text. PM: PubMed, EH: EvidenceHunt.

| Cochrane review | Systemic review title | Population text | Intervention text | PM hits (N) | EH hits (N) | N trials avail. | N trials identif. | % trials identif. |
| --- | --- | --- | --- | --- | --- | --- | --- | --- |
| 1, CD013739.pub0 | Prophylactic anticoagulants for people hospitalised with COVID-19 | Covid-19 | Antithrombin Dabigatran<br>Anticoagulant<br>anticoagulation<br>antithrombotic heparin<br>Fondaparinux Hirudin<br>Rivaroxaban Enoxaparin<br>reviparin Dalteparin<br>danaparoid edoxaban<br>Phenprocoumon Nadroparin<br>Acenocoumarol | 426 | 345 | 5 | 4 | 80% |
| 2, CD013740.pub2 | Mindfulness-based psychological interventions for improving mental well-being in medical students and junior doctors | student doctor<br>medical "house officer" resident<br>physician "medical school" graduate<br>undergraduate<br>postgraduate intern | mindfulness relaxation<br>MBCT "meditation" "mind training" | 1448 | 387 | 12 | 12 | 100% |
| 3, CD013750.pub0 | Platinum-containing regimens for triple-negative metastatic breast cancer | breast cancer | platinum cisplatin carboplatin | 898 | 771 | 9 | 9 | 100% |
| 5, CD013756.pub2 | Implanted spinal neuromodulation interventions for chronic pain in adults | <none> | "spinal cord stimulation"<br>"spinal cord electrostimulation" "dorsal root stimulation" "dorsal root electrostimulation" "epidural stimulation" "epidural electrostimulation"<br>neurostimulation SENZA neuromodulation | 2663 | 872 | 23 | 21 | 92% |
| 6, CD013757.pub2 | Palivizumab for preventing severe respiratory syncytial virus (RSV) infection in children | <none> | palivizumab | 230 | 223 | 10 | 8 | 80% |
| 7, CD013761.pub2 | Beds, overlays and mattresses for preventing and treating pressure ulcers: an overview of Cochrane Reviews and network meta-analysis | "pressure ulcers" | <none> | 1133 | 252 | 6 | 6 | 100% |
| 8, CD013776.pub2 | Blue versus white light for transurethral resection of non-muscle invasive bladder cancer | "Bladder cancer" | hexaminolevulinic fluorescent HAL light | 6174 | 453 | 24 | 22 | 92% |
| 12, CD013790.pub2 | Heated tobacco products for smoking cessation and reducing smoking prevalence | <none> | Carbon-Heated "heated tobacco" "tobacco heating" "heated cigarette" | 1756 | 300 | 16 | 14 | 88% |
| 13,<br>CD013792.pub2 | Progestogens for preventing miscarriage: a network meta-analysis | abortion miscarriage fetal pregnancy | progesterone dydrogesterone progestin Hydroxyprogesterone Medroxyprogesterone | 2826 | 1278 | 10 | 9 | 90% |
| 17,<br>CD013814.pub0 | Acupuncture for chronic nonspecific low back pain | "back pain" | acupuncture | 342 | 189 | 25 | 22 | 88% |
| 19,<br>CD013825.pub2 | SARS-CoV-2-neutralising monoclonal antibodies for treatment of COVID-19 | coronavirus COVID-19 "Sars-cov-2" | antibody Bamlanivimab Etesevimab Casirivimab imdevimab | 2553 | 529 | 7 | 7 | 100% |
| 20,<br>CD013836.pub2 | Antibiotic regimens for late-onset neonatal sepsis | neonatal sepsis | antibiotic vancomycin cefazolin meropenem ampicillin gentamicin amikacin | 2259 | 214 | 5 | 4 | 80% |
| 21,<br>CD013837.pub2 | Antibiotic regimens for early-onset neonatal sepsis | neonatal sepsis | antibiotic vancomycin cefazolin meropenem ampicillin gentamicin amikacin | 2259 | 214 | 7 | 5 | 71% |
| 22,<br>CD013839.pub2 | Through-knee versus above-knee amputation for vascular and non-vascular major lower limb amputations | <none> | amputation | 4929 | 9775 | 5 | 4 | 80% |
| 25,<br>CD013870.pub2 | Phototherapy for atopic eczema | eczema dermatitis | phototherapy ultraviolet UVB | 2801 | 354 | 27 | 21 | 78% |
| 26,<br>CD013874.pub2 | Rituximab for people with multiple sclerosis | multiple sclerosis ms "disseminated sclerosis" | rituximab rtx | 352 | 169 | 9 | 7 | 78% |
| 30,<br>CD013881.pub0 | Interleukin-6 blocking agents for treating COVID-19: a living systematic review | covid corona sars-cov-2 | interleukin-6 tocilizumab | 865 | 957 | 2 | 2 | 100% |
| 31,<br>CD014201.pub0 | Clotting factor concentrates for preventing bleeding and bleeding-related complications in previously treated individuals with haemophilia A or B | clotting factor hemophilia haemophilia "bleeding-related complication" | recombinant FVIII nonacog | 5181 | 793 | 17 | 15 | 88% |
| 32,<br>CD014484.pub0 | Low-dose oral misoprostol for induction of labour | labor labour birth induction | misoprostol | 872 | 769 | 32 | 31 | 97% |
| 38,<br>CD014739.pub0 | Antithrombotic therapy for ambulatory patients with multiple myeloma receiving immunomodulatory agents | myeloma "immunomodulatory agents" | antithrombotic enoxaparin | 15 | 5 | 1 | 1 | 100% |
| 40,<br>CD014953.pub2 | Virtual reality training for cataract surgery operating performance in ophthalmology trainees | cataract ophthalmological ophthalmology | simulator VR "virtual reality" "VR training" | 107 | 95 | 4 | 4 | 100% |
| 41,<br>CD014955.pub0 | Day care as a strategy for drowning prevention in children under 6 years of age in low- and middle-income countries | drowning | prevention | 155 | 40 | 2 | 1 | 50% |
| 42,<br>CD014962.pub0 | Remdesivir for the treatment of COVID-19 | covid corona sars-cov-2 | remdesivir | 548 | 269 | 3 | 2 | 67% |
| 43,<br>CD014963.pub0 | Systemic corticosteroids for the treatment of COVID-19 | covid corona sars-cov-2 | systemic corticosteroids methylprednisolone | 1696 | 585 | 3 | 1 | 33% |
| 44,<br>CD015017.pub2 | Ivermectin for preventing and treating COVID-19 | covid corona sars-cov-2 | ivermectin mectizan stromectol antiparasitic | 194 | 153 | 7 | 6 | 86% |
| 45,<br>CD015025.pub0 | Antibiotics for the treatment of COVID-19 | covid corona sars-cov-2 | anti-infective "anti-biotic" "anti-microbial" antibiotic antimicrobial azithromycin doxycycline | 708 | 2055 | 3 | 3 | 100% |
| 46,<br>CD015043.pub0 | Vitamin D supplementation for the treatment of COVID-19: a living systematic review | covid corona sars-cov-2 | vitamin D "vitamin D3" cholecalciferol ergocalciferol | 2456 | 1851 | 2 | 2 | 100% |
| 47,<br>CD015045.pub0 | Colchicine for the treatment of COVID-19 | covid corona sars-cov-2 | colchicine | 153 | 117 | 2 | 2 | 100% |
| 48,<br>CD015061.pub0 | Interventions for palliative symptom control in COVID-19 patients | covid corona sars-cov-2 | hospice palliative rehabilitation acupressure | 897 | 2558 | 3 | 0 | 0% |
| 50,<br>CD015374.pub0 | Chemotherapy for second-stage human African trypanosomiasis: drugs in use | trypanosomiasis sleeping | chemotherapy anti-trypanosomal fexinidazole acoziborole nifurtimox eflornithine | 1006 | 600 | 1 | 1 | 100% |

The number of hits returned from the search varied considerably, with a minimum of five and a maximum of 9775 (**Table 2**). In most searches (i.e., 75%), the number was below 788 (median: 371).

The average number of hits returned from PubMed was significantly higher (median 952; **Table 2**): Compared to the new approach, twenty-six of the thirty reviews returned higher volumes on PubMed (proportion: 0.87, 95% confidence interval: 0.69–0.96, p < 0.0001; exact binomial test).

## 4. Discussion

This study shows that an additional software layer between the user and the PubMed database can simplify and speed up identifying relevant clinical trials. With a straightforward set of keywords related to patient population and type of intervention, combined with PICO-text identification and a specific and sensitive study filter, key trials can be retrospectively identified as used in Cochrane systematic reviews. This platform does not replace the manual search and quality characterization of trials for inclusion in aggregated analyses. Still, it does offer the potential to bring current clinical evidence up to date much faster than usual. We consider our extension a valuable technology to keep up with the rapid growth of medical trial reports. The acquisition of relevant evidence in making scientific progress and managing health-related decisions is becoming increasingly difficult due to the large volume of published trials.

Freely assessable Internet platforms for helping users quickly and efficiently search and retrieve relevant publications from PubMed have been presented before. Wang et al. provided with their iPubMed platform a fuzzy search and autocomplete function (Wang et al., 2010). When their free platform was still online,^11^ they incorporated a software layer in front of PubMed, providing interactive feedback during query typing and approximate searching, allowing for minor spelling errors. The HubMed interface is dynamic and intuitive and integrates PubMed abstracts with data from other sources, including full-text links.^12^ Full-text links are provided through PubMed’s ELink service that leads to the document on the publisher’s website; via Ex Libris’ demonstration SFX server that offers a range of alternate full-text services through Google Scholar or a proxy server working with OpenURL linking standard (Eaton, 2006). The PubGet service (last available in 2017) linked searches to the full-text contents of open-access PDFs, allowing the user to directly display the result in context, bypassing the PubMed website.^13^ BabelMeSH provides a multilanguage online search platform by offering the translation of MeSH terms in eleven languages, including Arabic, Chinese, Dutch, Japanese, Korean, and Spanish.^14^ The query is constructed in the language of choice and translated into English before searching PubMed. The initial accuracy of compound keyword translation was 68% for French, 60% for Spanish, and 51% for Portuguese (Liu et al., 2006). PICO Linguist is comparable in software architecture to the multilingual BabelMeSH.^15^ Instead, it has a single input box structuring the query format into Patient/Problem (P), Intervention (I), Comparison (C), and Outcome (O) input forms (Fontelo et al., 2007). A different approach is taken by askMEDLINE (Fontelo et al., 2005). This software handles queries in the form of free-text questions or medical phrases in natural language. Initially developed to parse PICO-related questions and extract the patient, intervention, comparison, and outcome form, it was later launched as a tool for non-expert medical information. The tool has a GSpell spelling checker and a filter option to select the publication type, including ‘Clinical Trial’ and ‘Randomized Controlled Trial’ based on PubMed’s study type categorization.^16^ Compared to these services, our platform is user-friendly, fast, allows automatic notification, and is more up-to-date with the latest text-mining algorithms and database handling.

Our study has limitations. 1) We validated our keyword combinations on a retrospective set of included trials in published Cochrane reviews. The trials excluded for aggregated analysis – for example, due to low study quality – were not considered. Nor did we characterize the detection of complementary studies and secondary literature reported in the Cochrane reviews. Our database includes all PubMed records, so we do not expect different behavior in excluded, lower-quality trials or related non-trial literature. However, that would require an additional and much larger manually-labeled validation set. 2) We focused on systematic reviews with an interventional, and hence PICO-structured, research question. More and more diagnostic and prognostic systematic reviews are published, including evidence from trial and non-trial reports. Identifying relevant studies will probably also work in these clinical domains. Still, we have not validated it due to different search requirements and the non-PICO structure of the corresponding queries in non-interventional reviews. 3) PubMed contains most of the international biomedical literature. Still, our software and validation do not include different non-free databases, including available Embase, Web of Science, and Scopus. We restricted our study to the (single) freely available PubMed database.

4) Our validation was limited due to trial identification based on abstract text only. Manual raters investigate the full text to decide whether or not to include a clinical trial report in the aggregated systematic review analysis. On the other hand, we relied on Patient and Intervention sentences available in the abstract—the non-structured nature of trial summaries results in missing information in some cases. Labeling the Patient and Intervention sentences in the full text would probably increase the identification yield. Building a full-text database is only possible for reports published under a permissive license. Fortunately, the free PubMed Central (PMC) archive of biomedical and life sciences journal literature is growing exponentially, with clinical reports and preprints made available under license terms that allow reuse. In the last twenty years, forty percent of clinical trial reports have been published on PubMed with the option to access the free full text.^17^ A future hybrid database may combine full-text – when available – with abstract-only texts to increase the identification yield.

Other future directions include an extension of the PICO text labeling with NER models suitable for diagnostic and prognostic queries. Although our database is synchronized with PubMed daily, the updates are restricted to published trials only. We envision that rapid updates may include study identification within clinical trial registration registries (e.g., Clinical Trials (.gov),^18^ the EU Clinical Trials Register,^19^ and the Open Science Framework^20^) as more and more trials are documented before and during study conduct. These additional sources may help researchers and clinicians anticipate upcoming evidence. Equally interesting is combining the peer-reviewed PubMed trials with the not-yet peer-reviewed preprint trial reports – to enhance early trial identification – available in the preprint databases, including bioRxiv,^21^ PsyArXiv,^22^ medRxiv,^23^ or JMIR Preprints.^24^

More rigorous search and database transformations are within reach with the latest development of large-language models (Singhal et al., 2023). Medical text models, like the generative Med-PaLM-2, perform on text comprehension encouragingly well, answering Medical Licensing Examination (USMLE)-style questions with 85% accuracy and passing the MedMCQA medical examination dataset questions with a 72.3% score.^25^ As soon as language models can reliably understand and retrieve Patient and Intervention sentences from the full PubMed database, the research question may replace the query and yet identify the relevant clinical trial reports. Open-source language models like MedAlpaca are worth investigating if effectively coupled with the PubMed database and a user-friendly ‘chat’ platform (Han et al., 2023).

## Data Availability

All data produced in the present study are available upon reasonable request to the authors

https://evidencehunt.com/browse

## Footnotes

1 https://www.cochranelibrary.com/cdsr/about-cdsr

2 https://www.nlm.nih.gov/medline

3 ftp://ftp.ncbi.nlm.nih.gov/pubmed/baseline

4 ftp://ftp.ncbi.nlm.nih.gov/pubmed/updatefiles

5 https://evidencehunt.com/browse/

6 PubMed query: all[sb]

7 https://www.elastic.co/guide/en/enterprise-search/current/start.html

8 https://doccano.herokuapp.com

9 https://huggingface.co/microsoft/BiomedNLP-PubMedBERT-large-uncased-abstract

10 That is: ( <P terms>[mh]/[tiab] ) AND ( <I terms>[mh]/[tiab] ) AND ( ( "randomized controlled trial"[pt] OR "controlled clinical trial"[pt] OR randomized[tiab] OR placebo[tiab] ОR "drug therapy"[sh] OR randomly[tiab] OR trial[tiab] OR groups[tiab] ) NOT (animals[mh] NOT humans[mh] ) ), The Cochrane Collaboration. ‘Cochrane Handbook for Systematic Reviews of Interventions Version 6.3’, Technical Supplement to Chapter 4: Searching for and selecting studies, 2022.

11 http://ipubmed.ics.uci.edu and http://tastier.cs.tsinghua.edu.cn/ipubmed

12 https://www.hubmed.org

13 https://en.wikipedia.org/wiki/Pubget

14 http://babelmesh.nlm.nih.gov

15 https://pubmedhh.nlm.nih.gov/pico/consensus.php

16 https://pubmedhh.nlm.nih.gov/ask/index.php

17 Based on the hits from the query all[sb] restricted to the time period 2004-2024 and article type filter ‘Clinical Trial’ divided by the hits from the same query with no article type filter activated: 242k / 599k = 40.4% (Aug 14, 2023).

18 https://classic.clinicaltrials.gov

19 https://www.clinicaltrialsregister.eu

20 https://osf.io

21 https://www.biorxiv.org

22 https://psyarxiv.com

23 https://www.medrxiv.org

24 https://preprints.jmir.org

25 https://blog.google/technology/health/ai-llm-medpalm-research-thecheckup/

